# Sex Disparities in Cognitive Impairment Research: A Scoping Review in Informatics Literature

**DOI:** 10.1101/2024.12.27.24319704

**Authors:** Muskan Garg, Xingyi Liu, Jie Lin, Maria Vassilaki, Ronald C Petersen, Jennifer St Sauver, Ekta Kapoor, Sunghwan Sohn

## Abstract

**Rationale:** A scoping review was conducted to investigate knowledge gaps in the informatics research literature regarding sex differences in cognitive decline, identifying existing studies and areas where further studies are needed.

**Materials and Methods:** We searched Ovid and other databases for studies on sex differences and cognitive decline, focusing on publications in peer-reviewed informatics journals and conference proceedings from 2000 to 2023. The selected manuscripts were analyzed and summarized through discussion among three reviewers.

**Results:** A total of 13 articles were selected and examined for metadata and attributes analysis. Most studies are conducted in United States (n=5) and European Union (n=4), about a half are published after 2020 (n=6), and most studies are published in Springer and Elsevier. Our attributes-based analysis highlights the different aspects of reported studies such as task, method, dataset and its size, and sex-specific inferences.

**Discussion:** Sex-specific disparities in cognitive decline remain a critical issue in healthcare, yet most informatics research has primarily concentrated on identifying basic sex differences, such as tracking the progression of cognitive decline in men and women. While these studies are valuable, they fall short of addressing the more complex underlying causes of these sex-specific disparities in progression of cognitive decline.

**Conclusion:** There is a significant gap using informatics in understanding how biological, social, and behavioral factors contribute to sex-specific disparities. This limited focus restricts the development of effective intervention strategies for mitigating sex-specific differences in cognitive health outcomes, underscoring the need for more comprehensive research that goes beyond mere identification to find the root cause of these disparities in healthcare.

## 1. Introduction

Around 6.2 million Americans aged 65 and above are diagnosed with Alzheimer’s dementia. Without significant medical advancements that can prevent, halt, or cure Alzheimer’s Disease (AD), this figure might double, reaching 13.8 million by 2060 [1, 2]. Existing studies in healthcare informatics demonstrate significant impacts on the quality of life due to cognitive decline and present substantial challenges to healthcare systems [3]. Personalized medicine is becoming increasingly important in healthcare as the prevalence and progression of cognitive decline appear to differ between men and women [4]. Identifying sex-specific trends in cognitive decline can help healthcare providers anticipate and allocate resources more efficiently [5, 6]. By understanding how these impacts differ between men and women, healthcare providers can offer more supportive and effective care, improving the overall quality of life for all patients through better policymaking and the adoption of more effective preventive strategies.

Most existing studies have focused on identifying basic sex differences, such as variations in the diagnosis, progression, and treatments of cognitive decline, without carrying out in-depth informatics-based analysis. A cohort study suggests women may have greater cognitive reserve (global cognition, executive function, and memory) but experience faster cognitive decline than men, contributing to sex differences in late-life dementia risk [7]. Such sex-specific health disparities within healthcare informatics constitute a crucial and concerning area of investigation for the informatics research community.

Despite the significant scientific advances achieved so far, most of the currently used healthcare informatics methods do not account for bias detection [6, 8, 9]. Most study designs in informatics literature lack consideration of the sex dimensions, thereby overlooking their impact on advanced cognitive decline differences among participants such as causal analysis [10, 11]. Evidence-based decisions for sex-specific cognitive decline may allow healthcare informatics to better resource allocation, improved precision medicine, real-time monitoring of health outcomes. This personalized approach can enhance patient outcomes, reduce the risk of misdiagnosis, and ensure that both men and women receive care that is optimally suited to their unique needs.

We investigated the existing literature and identified surveys and reviews that address sex-specific disparities in cognitive decline. Past reviews investigated evidence on cognitive impairment differences by sexual orientation from clinical studies [12, 13]. While existing surveys and reviews primarily focus on clinical trials, there is a notable gap in investigating the informatics literature-based study designs for discovering sex differences in cognitive decline.

Our scoping review aims to examine the existing informatics literature on sex-specific disparities for cognitive decline. We seek to identify and categorize existing studies to understand the breadth and depth of current research. We highlight areas where informatics has successfully contributed to understanding these disparities and identify gaps where further research is needed.

Our paper is organized in different sections. Section 2 outlines the databases and search strategy employed to compile a list of published articles. We describe the inclusion and exclusion criteria applied and conclude with an analysis of the content of the selected articles. In Section 3, we discuss the metadata statistics and attributes of the reported studies. We frame a list of themes and sub-themes to analyze the existing literature. In Section 4, we discuss the summary of our key findings and limitations of existing literature. We conclude with future scope in Section 5.

## 2. Materials and Methods

### 2.1. Bibliography Search

The scoping review targeted publications on sex-specific differences in peer-reviewed informatics journals and conference proceedings published between 2000 and 2023 in English language. Based on this criterion, along with the inclusion and exclusion principle, a senior librarian devised a searching strategy for identifying articles from a set of databases (see Table 1). The search criteria of article selection are depicted in Figure 1.

**Figure 1:**
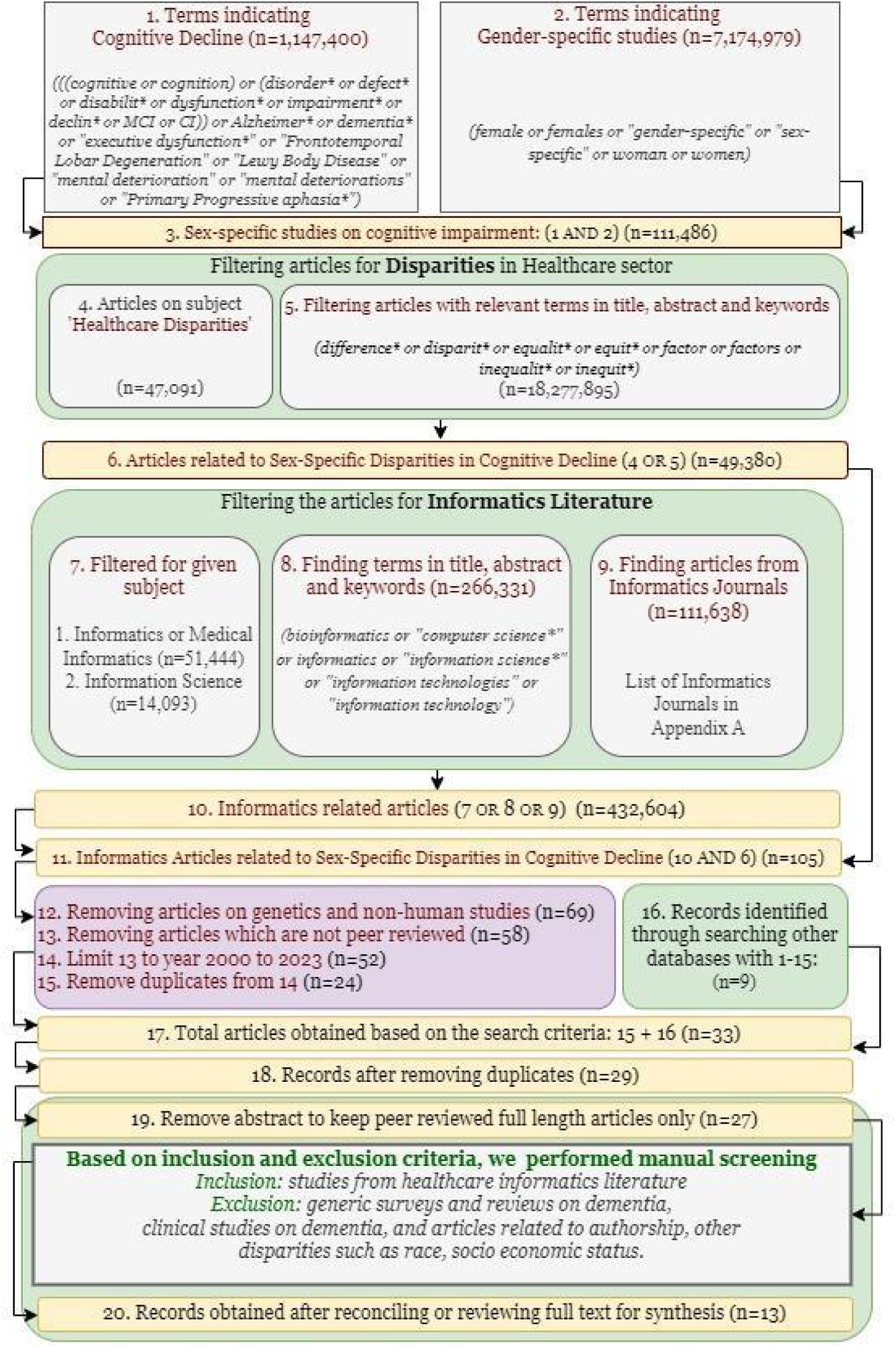
Overview of the search strategy used for selecting articles. See List of Informatics Articles in Appendix A, and terms related to articles on genetics, non-human studies, and non-peer reviewed venues in Appendix B. A full checklist of the preferred reported items for PRISMA extension of scoping reviews (PRISMA-ScR) [6] is available in Appendix C.

**Table 1:** List of Databases, used as a source of article collection.

| **Database(s)** | **Coverage Period** |
| --- | --- |
| APA PsycInfo | 1806 to April, 2024 |
| EBM Reviews - Cochrane Central Register of Controlled Trials | Till March 2024 |
| EBM Reviews - Cochrane Database of Systematic Reviews | 2005 to April, 2024 |
| Embase | 1974 to April, 2024 |
| Ovid MEDLINE(R) and Epub Ahead of Print, In-Process, In-Data-Review & Other Non-Indexed Citations, Daily and Versions | 1946 to April, 2024 |

## 3. Results

We present the results in two parts: (i) the metadata analysis to examine the bibliographic landscape of the research field and its development over time, (ii) the attributes of the studies (clinical condition, methodology, and study data) to assess the reliability and validity of the findings and understand the context in which the studies were conducted. We provide discussion to reveal gaps in the literature, highlight areas of consensus and debate, and suggest directions for future research.

### 3.1. Metadata Statistics

We analyzed the bibliometric information to identify research trends, such as publication country, publisher, and publication year (Figure 3). More than 35% (n=5) publications are from the USA, followed by the European Union (n=4). About half of the eligible studies are published in or after 2020 (n=6). The sex-specific disparities have recently gained pace in 2023 (see Figure 3(b)). Almost half of the studies were published by JMIR publications and Springer (n=6), followed by Wiley and Elsevier as shown in Figure 3(c).

**Figure 3:**
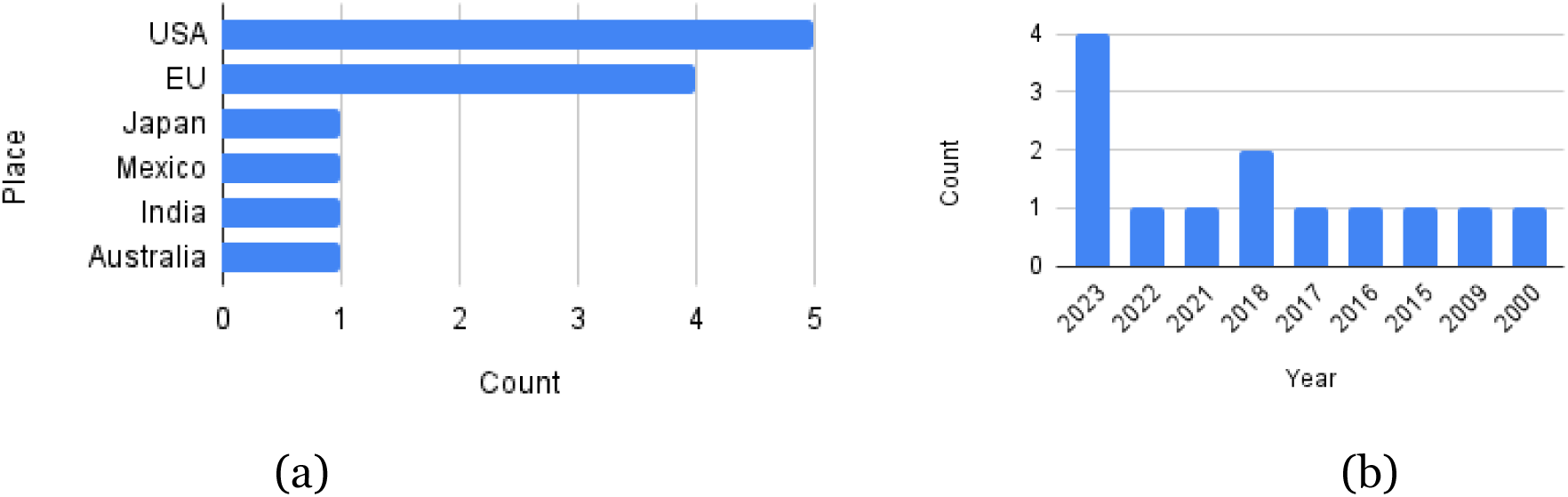
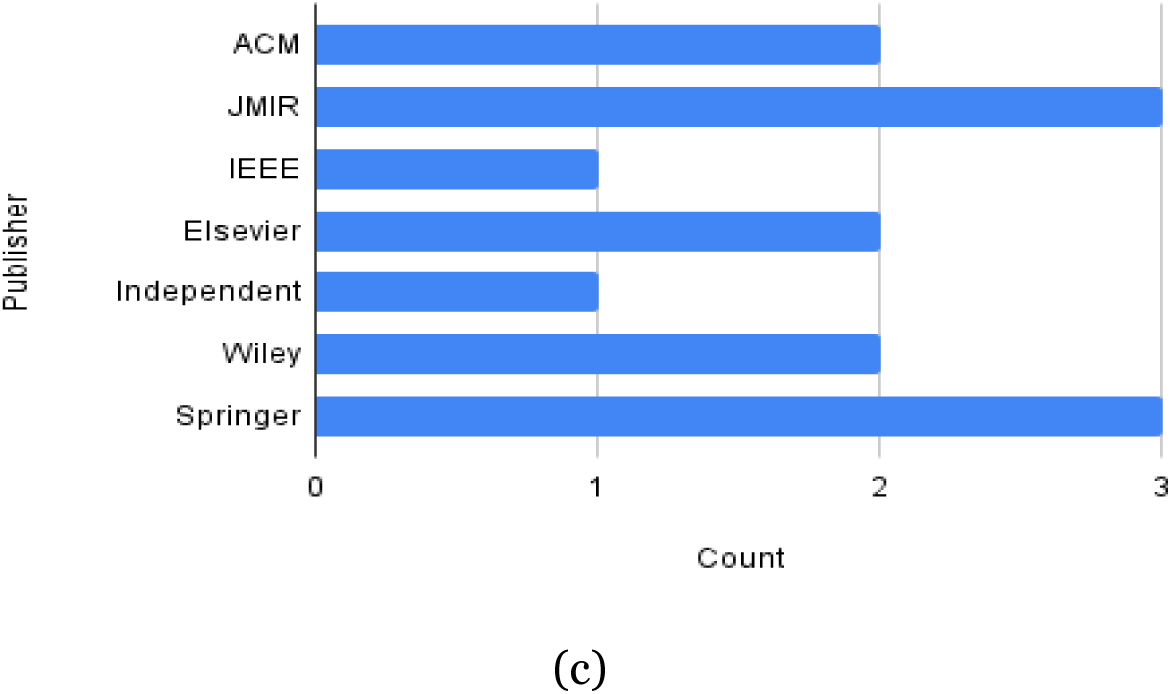
Distribution of publication in terms of their (a) place, (b) year, and (c) publisher.

### 3.2. Attributes of reported studies

We explored four key attributes, such as clinical condition, methodology, and study data size, to comprehensively understand the research landscape related to sex-specific disparities. Table 1 provides a summary of the attributes of the studies we reported. Other than brief information about these attributes, we investigate each of these attributes such as study title, clinical conditions, clinical tasks, study size, the role of informatics in detail (see Appendix D).

#### 3.2.1. Sex-specific analysis

Research on sex-specific differences in cognitive decline has highlighted distinct patterns across multiple dimensions. Women have been found to exhibit more severe cognitive impairment and experience a faster rate of cognitive decline compared to men at the onset of Alzheimer’s disease (AD) [14]. Sociodemographic factors, such as lower education levels and socioeconomic status, disproportionately affect older women, increasing their vulnerability to dementia [15–17]. Imaging studies have revealed sex-specific differences in brain metabolism, particularly in the thalamus and cerebellum, supporting biological distinctions in disease progression [15, 18, 19]. Furthermore, clustering analyses of AD patient populations demonstrate divergent patterns, with women forming fewer but more defined clusters compared to men, indicating potential variability in disease trajectories [16].

Predictive models for cognitive decline show comparable performance across sexes in some frameworks [19, 20], such as elastic net regression, though men tend to exhibit higher c-statistic values in Cox regression analyses. In terms of healthcare utilization, men with AD are more likely to be hospitalized in their final year of life, while women with cognitive decline, especially those with comorbidities and lower physical function, are more likely to rely on assistive technology and telecare services. Moreover, women with dementia are less likely to participate in clinical studies, despite being slightly overrepresented in some cohorts, such as the Health eHeart study[21].

Caregiver studies reflect the gendered dynamics in dementia care, with a higher proportion of female caregivers, particularly in Hispanic rural populations [22, 23]. Women with dementia are also at increased risk for long-term care needs, and their caregivers—primarily female spouses and daughters—often express varied emotional responses to caregiving programs [24]. Collectively, these findings emphasize the importance of considering sex-specific factors in cognitive decline research, with implications for tailored interventions, healthcare delivery, and support mechanisms across different populations.

#### 3.2.2. Approaches

This section summarizes the approaches in reviewed articles to improve the understanding, diagnosis, and management of AD, dementia, and other related conditions, with particular emphasis on addressing sex-specific disparities. Among 13 articles, 5 are associated with *public health*. From encompassing the need of care for individuals with dementia [22, 24, 25] to behavioral analysis with the decision making [21, 23], the informatics literature has contributed toward better treatment plans for people with dementia. In their efforts, they draw attention toward the equitable care-need levels by caregivers, nursing, medical, and allied health services. They emphasize the need of an in-depth analysis for the difference in help-seeking behaviors and the decision making among men and women.

We also identified studies using descriptive analysis [16, 17] and predictive analytics [14, 20] for individuals with mild cognitive impairment (MCI) or dementia. Following their examination of cohort characterization and dementia prediction, the studies underscore the need for evidence-based quantification of sex-specific disparities in healthcare. While identifying risk factors associated with cognitive decline has gained significant attention in recent years, only two studies have specifically focused on informatics-based analyses of risk factors for cognitive decline in men and women [18, 26]. We also observed the technological advancements supporting neuropsychological tests [19] and telecare for home-dwelling individuals with dementia [15]. These findings highlight the unmet potential for reliable communication and sensing technologies for this population.

#### 3.2.3. Study data

The reviewed studies used various types of data including multisite study data (Alzheimer’s Disease Neuroimaging Initiative [ADNI]) which is an open-access series of imaging and clinical data designed to enhance research into AD diagnosis and progression [14, 16, 18]. The dataset includes various imaging modalities, such as structural MRI, functional MRI (fMRI), and PET scans, alongside cerebrospinal fluid (CSF) biomarkers and comprehensive neuropsychological test scores [27, 28]. Analytical techniques are used with moderate sample sizes to enable subgroup analysis based on sex and race [18]. Risk factor analysis, descriptive and predictive modeling studies utilize larger datasets, divided into training and test sets, to enhance the robustness of the models and their applicability to diverse populations [14, 16, 19, 20, 24, 26]. Recruitment studies often involve extensive data collection efforts, exemplified by large-scale email campaigns that engage thousands of participants [21]. Observational analysis with large datasets enhances the reliability of evidence-based findings in informatics literature. Nevertheless, establishing frameworks for sex-specific analyses can also be effectively achieved with smaller datasets, contingent upon the study design.

**Table 1:**
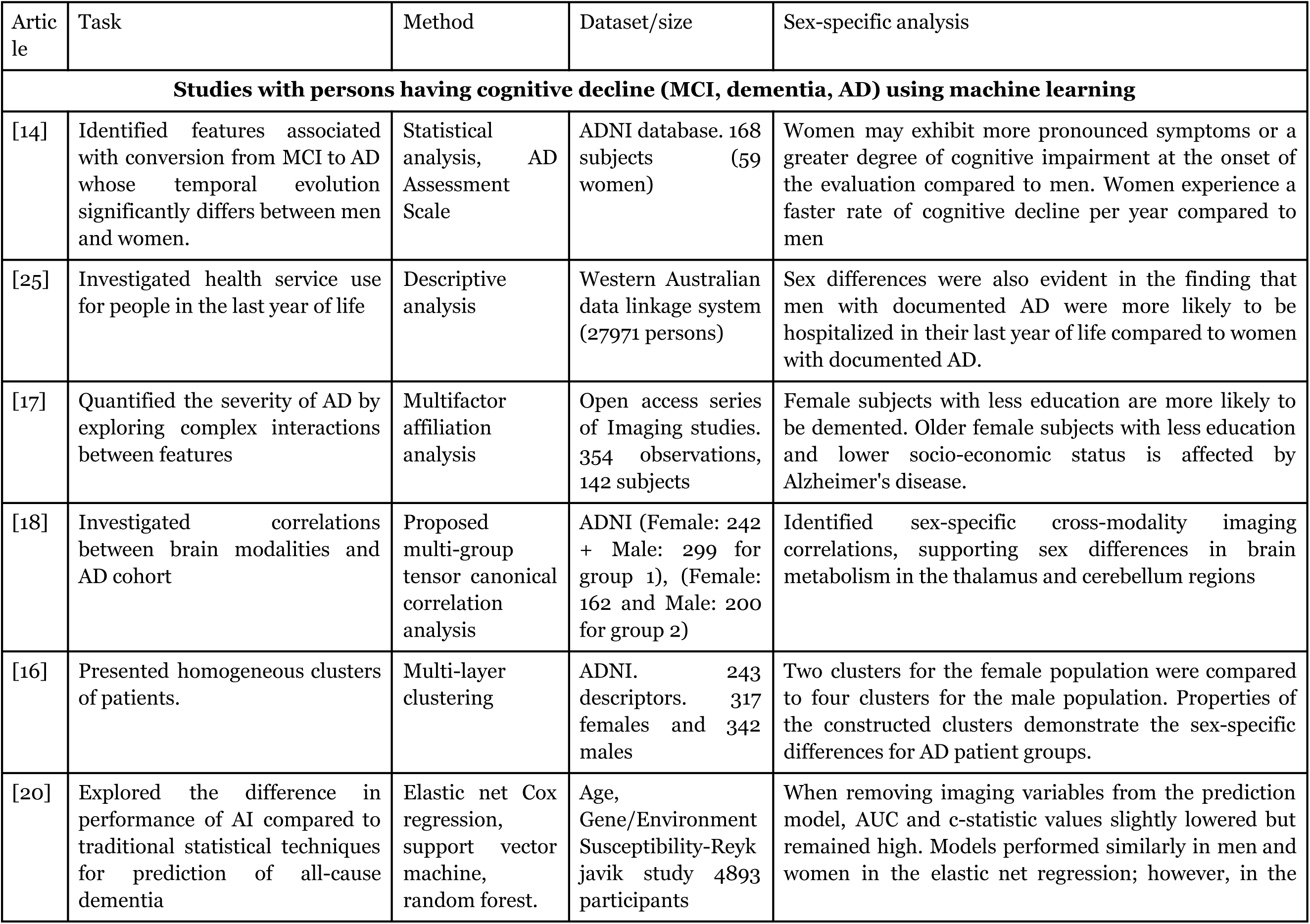
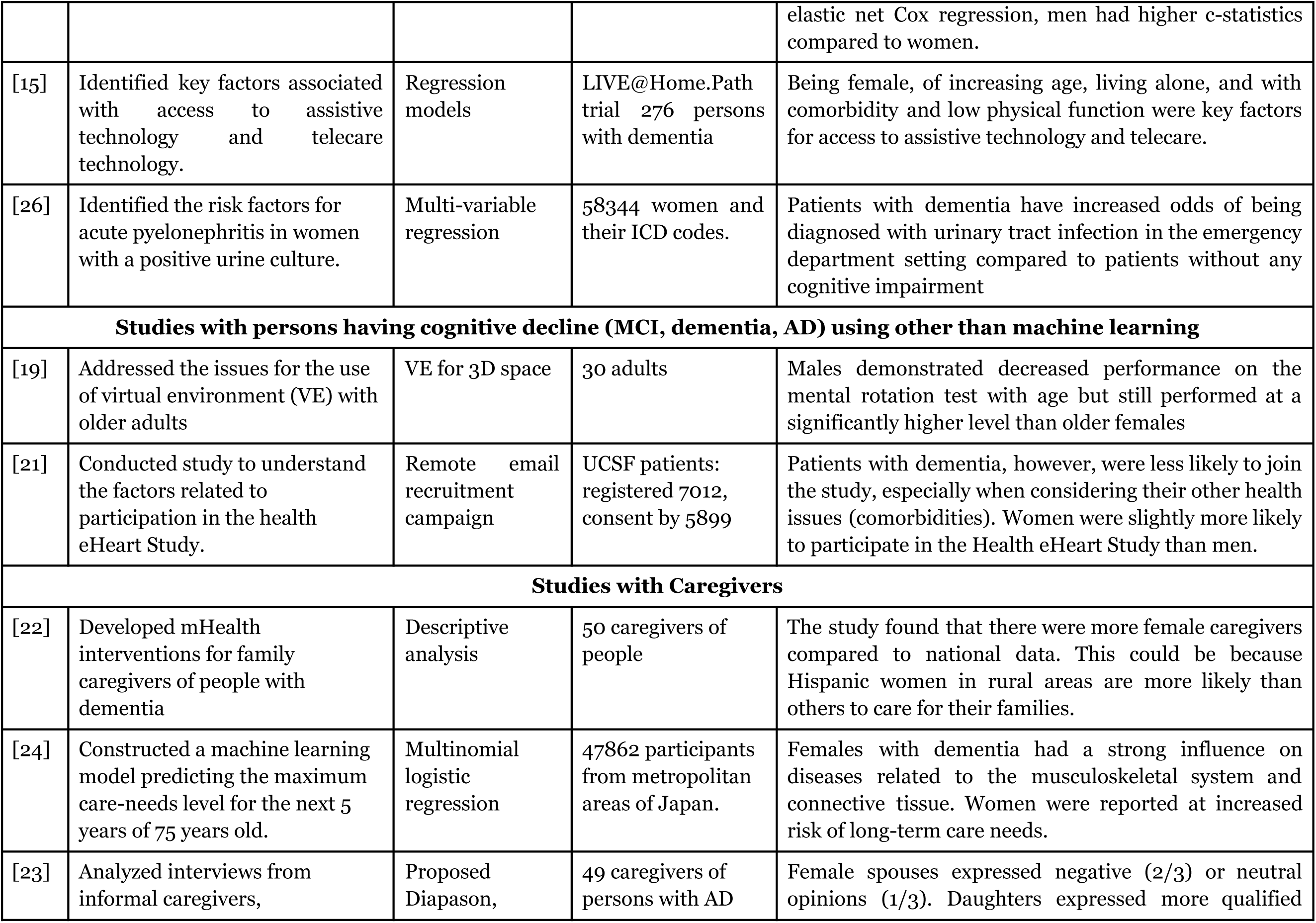
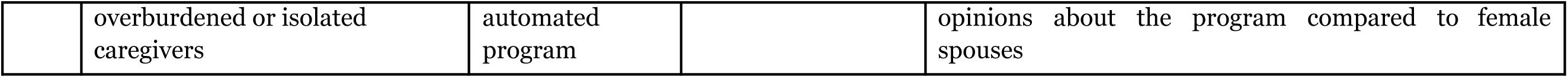
Summary of attributes of reported studies.

### 3.3. Thematic analysis of reported studies

Inferences drawn from these studies indicate that women and men may experience different trajectories of cognitive decline. To obtain a clear picture of how current research in healthcare informatics intersects with studies on sex-specific differences, we classified the articles thematically in Table 2.

**Table 2:** Thematic classification of articles for healthcare informatics and sex-specific differences.

|  | [18] | [17] | [23] | [25] | [19] | [20] | [21] | [24] | [26] | [22] | [15<br>] | [14] | [16] |
| --- | --- | --- | --- | --- | --- | --- | --- | --- | --- | --- | --- | --- | --- |
| Healthcare Informatics |  |  |  |  |  |  |  |  |  |  |  |  |  |
| CDSS |  | X |  |  |  |  | X | X | X |  | X | X | X |
| AI | X |  |  | X | X | X |  | X |  |  |  |  | X |
| TTM |  |  | X |  |  |  |  |  |  | X | X |  |  |
| Sex-specific differences |  |  |  |  |  |  |  |  |  |  |  |  |  |
| S-PA |  |  |  |  | X | X |  | X | X | X |  | X | X |
| S-PN | X | X |  |  |  |  |  |  |  |  |  | X | X |
| S-DP |  |  | X | X |  |  | X |  |  |  | X |  |  |

TTM: Telemedicine, Telehealth, and Mobile Health (mHealth) Apps; CDSS: Clinical Decision Support Systems; AI: Artificial Intelligence, health analytics, big data; S-PA: Sex Differences in Cognitive decline patterns and Assessment; S-PN: Sex Differences in Pharmacological and Neuroimaging studies; S-DP: Sex Differences in Digital health, behavioral, and Psychosocial, demographic Factors.

Clinical Decision Support Systems (CDSS) is being actively explored, particularly in terms of how it can integrate with other advanced technologies like AI and health analytics [29]. The interest in CDSS suggests a growing recognition of the need for systems that can assist clinicians in making better-informed decisions, potentially considering sex-specific differences. AI is a major focus area, particularly in its potential to revolutionize healthcare through predictive analytics, personalized treatment, and big data analysis. The frequent intersection with CDSS suggests that AI is increasingly being integrated into decision-support frameworks. The focus on Telemedicine, Telehealth, and Mobile Health (mHealth) Apps (TTM) indicates a shift towards digital health solutions, especially important in a post-pandemic world and mobility impairment of elderly persons where remote healthcare has become essential [30]. The integration of TTM with sex-specific differences research suggests that digital health platforms are being studied for their effectiveness across different sex demographics.

There is a significant research focus on how cognitive decline differs between sexes, which is critical for developing sex-specific interventions and treatment protocols. Studies on how pharmacological treatments and neuroimaging results differ between sexes could lead to more effective and personalized drug therapies and a better understanding of neurodegenerative diseases. Research in this area highlights the importance of considering sex differences in digital health adoption, user behavior, and psychosocial factors.

## 4. Discussion

In this section, we discuss the summary of the reviewed articles, our inferences and limitations of this study. Our thematic analysis indicates that exploring hormonal influences, genetic biomarkers, and structural differences in the brains of men and women could reveal significant insights into sex-specific disparities in dementia [31, 32]. Additionally, investigating how lifestyle factors, environmental influences, and genetic predispositions affect the onset and progression of dementia may deepen our understanding of these differences. Furthermore, considering how sex-specific disparities intersect with other demographic factors—such as race, ethnicity, and socio-economic status—can help elucidate the compounded effects on dementia risk and progression [33]. This holistic approach is crucial for developing more effective, inclusive, and tailored intervention strategies for personalized treatment plans. Advancing and evaluating technologies like wearable devices and mobile health applications could enhance the monitoring and management of Alzheimer’s disease and dementia in a manner that accounts for sex-specific differences.

Considering these observations, we propose several directions for future research. These recommendations are informed by current gaps in the literature and emerging trends in the field. First, designing and testing interventions that are specifically tailored to the needs of men and women is crucial for optimizing treatment outcomes and enhancing patient care. Second, conducting long-term, large-scale longitudinal studies will be invaluable in tracking sex-specific disease trajectories from early stages of cognitive decline through advanced dementia. These studies can help identify critical windows for intervention and prevention. Third, utilizing advanced data analytics and machine learning techniques is also recommended to analyze large and complex datasets. These methods can uncover patterns and predictors that traditional approaches may overlook, potentially leading to new insights and advancements in dementia research. Finally, the integration of various data sources—such as electronic health records, wearable devices, and patient-reported outcomes—will provide a comprehensive view of patients’ health and cognitive status.

Our search strategy was limited to informatics literature for sex-specific disparities in cognitive decline and might not have captured all relevant studies due to variations in terminology. Informatics interacts with the field of sex-specific disparities in multiple ways, so we focused on subject-specific literature, search terms in abstracts, and publications in informatics journals in OVID database. Additionally, as the discipline of informatics has diversified in its publication avenues, primary informatics journals and conferences alone may not fully represent its scope. We acknowledge that our synthesis may be influenced by our subjective interpretations, yet we strive to draw inferences about various aspects of each theme.

## 5. Conclusion

This scoping review summarizes the utilization of healthcare informatics, particularly in neuroimaging, machine learning applications, and cognitive assessment tools, underscoring their potential to improve early diagnosis and management of neurodegenerative diseases like Alzheimer’s. It reveals critical insights into sex-specific disparities in healthcare, emphasizing the need for targeted interventions and equitable access to resources. The review identifies different themes in predicting cognitive decline and developing effective prevention and intervention strategies with healthcare informatics and sex-specific differences, in the reported studies. Despite the increasing use of informatics in the field, challenges such as data quality, model interpretability, and technology adoption persist, underscoring the necessity for further research, validation, and comprehensive approaches to enhance healthcare delivery and outcomes for diverse populations.

## Data Availability

No data is used for scoping review

https://github.com/drmuskangarg/scoping_review_sex_disparity

## DECLARATIONS

### Ethics approval and consent to participate

This study was approved by the Mayo Clinic Institutional Review Board and the Olmsted Medical Center Institutional Review Boards. No consent is required for this scoping review.

### Consent for publication

No consent is required for publication of this study.

### Availability of data and materials

Appendices include titles of all journals and conference proceedings covered by the search (Appendix A), terms for search strategy (Appendix B), details of selected studies (Appendix C), attributes (Appendix D) available in our GitHub repository^1^.

### Authors’ contributions

M.G. conceptualized and designed the study, collected, analyzed, and interpreted the data, drafted the initial manuscript, and revised the manuscript. M.V. and J.S. analyzed and interpreted the data and revised the manuscript. X.L., J.L., E.K., and R.P. interpreted the data and revised the manuscript. S.S conceptualized and designed the study, supervised data collection and analysis, interpreted the data, and reviewed the manuscript. All authors approved the final manuscript and agreed to be accountable for all aspects of the work.

## Acknowledgements

We acknowledge that Prokop, Larry J., a senior librarian, acquired a list of publications based on the given criteria for scoping review.

## Footnotes

1 https://github.com/drmuskangarg/scoping_review_sex_disparity

